# Practitioner-Related Factors Influencing Pediatric Contact Lens Fitting for Myopia Management in Kenya: A Cross-Sectional Survey

**DOI:** 10.64898/2025.11.30.25341304

**Authors:** Gellause Murefu Kololi, Tecla Jerotich Sum

## Abstract

**Objective:** To examine practitioner-related factors influencing pediatric contact lens (CL) fitting for myopia management in Kenya and to identify key barriers and facilitators shaping prescribing practices.

**Methods and Analysis:** This cross-sectional survey involved 148 eye care practitioners (ECPs) from 38 clinics. A validated questionnaire captured demographics, clinical decision-making, and perceived enablers or barriers to pediatric CL prescribing. Data were analyzed using descriptive statistics and multivariate logistic regression at p < 0.05.

**Results:** Most ECPs prescribed spectacles for children under 8 years (79.1%) and initiated CLs at 13–14 years. Major facilitators were child motivation (OR = 25.04, CI: 4.36–46.03, p = 0.01), trial lens availability (OR = 2.89, CI: 1.05–5.47, p = 0.02), and perceived market opportunity (OR = 8.74, CI: 4.32–14.70, p = 0.01). Barriers included low public awareness (OR = 0.14, CI: 0.05–0.32, p = 0.01) and practitioner inexperience (OR = 0.17, CI: 0.08–0.35, p = 0.01).

**Conclusions:** Pediatric CL prescribing in Kenya remains limited by low awareness and practitioner inexperience. Availability of trial lenses and motivated young patients enhance uptake. Targeted practitioner training and community education could improve early myopia-control practices and support wider adoption of pediatric CLs. However, interpretation of these findings is limited by the self-reported nature of the practitioner data.

**Key Message of This Article:** *What is already known on this topic:* Childhood myopia causes significant educational and social disadvantages. Despite its growing prevalence, pediatric contact lens prescribing remains limited in low-resource settings due to structural barriers, practitioner hesitancy, and parental concerns, while most existing evidence overlooks these systemic challenges in sub-Saharan Africa.

*What this study adds:* This study provides the first national-level analysis from Kenya quantifying how practitioner self-efficacy, parental attitudes, and trial-lens availability influence pediatric contact lens uptake. It applies a multivariate framework to isolate modifiable behavioral and structural predictors previously unexamined in this context.

*How this study might affect research, practice, or policy:* The findings generate a context-specific evidence base for advancing pediatric myopia-control programs through practitioner capacity-building, improved clinic resourcing, and integration of evidence-led policies within child eye health strategies.

## 1.0 Introduction

Myopia is a chronic, progressive eye disease that poses a global public health concern due to its impact on visual function, academic performance, and long-term ocular health—particularly in children and adolescents [1, 2]. Prevalence among school-aged children and young adults has reached up to 73% in East and Southeast Asia, with Europe and North America reporting rates between 20% and 40%[2]. In Africa, while early estimates placed childhood myopia at 5.5%[3], recent meta-analyses suggest a slightly lower average of 4.7% [4]. In Kenya, one study [5] estimated prevalence at 14% among school-aged children, highlighting the need for targeted interventions.

This rising trend places strain on Kenya’s already limited eye health infrastructure, where pediatric refraction, low-vision care, and affordable optical services remain inadequate [6]. These limitations not only threaten quality of life of school-going individuals with myopia but also widen disparities in education and public healthcare access [7].

Over recent decades, contact lenses (CLs) technology has greatly evolved, affecting its clinical application—including in pediatric refractive correction. An estimated 150 million people use contact lenses globally, with approximately 45 million users in the United States alone [8]. Data from the US. National Health Interview Survey indicate that 25.3% of children and adolescents aged 6–17 use corrective lenses, with higher use among girls (36.2%) [9].

Randomized trial studies show that orthokeratology and soft multifocal contact lenses slow myopia progression in children, while also reducing retinal image minification, widening the visual field, and improving clarity compared with conventional single-vision lenses [7, 10]. However, concerns remain, including infection risk and reduction of prismatic relief at near, which may increase accommodation and convergence demand during close work in some children [8].

Although attitudes toward pediatric contact lens fitting and use have become favorable, actual uptake has not risen in parallel [11, 12]. Despite notable advancements in contact lens design and materials, spectacles continue to dominate as the primary mode of refractive correction worldwide [3, 5, 12]. Persistent structural and behavioral barriers— such as cost, limited availability, parental concerns, and the absence of policy support— remain critical obstacles to wider CL adoption [13].

Eye care practitioners (ECPs) play a central role in facilitating contact lens uptake among children and adolescents. However, their attitudes and prescribing patterns vary considerably across regions. Global meta-analyses indicate that age remains a key criterion for many ECPs when determining eligibility for pediatric CL fitting [14, 15]. For instance, in Europe and the United States, pediatric CL prescribing is approached differently from adult prescriptions [14]. By contrast, the limited evidence from sub-Saharan Africa suggests that ECPs, in managing childhood myopia, often default to spectacles—reinforcing the gap between advances in myopia control research and the realities of eye care in low-resource settings [16, 17].

Factors such as comfort, oxygen permeability, and ease of handling can encourage CL prescribing, especially for motivated children [18]. However, little is known about how these competing practitioner barriers and motivators to uptake interact in low-resource contexts, leaving a critical gap in understanding what ultimately shapes pediatric CL fitting and uptake in sub-Saharan Africa.

In Kenya, CL services—mainly limited to soft lenses—are concentrated in urban clinics with minimal regulation. The lack of a national registry and continued reliance on spectacles highlight disparities in access [5]. Africa’s contact lens market may reach USD 62.6 million by 2029, yet Kenya’s USD 0.44 million share shows unmet need [19]. Data on availability, affordability, and appropriateness of pediatric CL services in Kenya remain scarce.

This study addresses the evidence gap by identifying barriers and enablers influencing contact-lens prescribing among Kenyan practitioners and assessing how these factors shape real-world uptake among myopic children—vital for improving pediatric vision and population eye health.

## 2.0 Materials and Methods

### 2.1 Study design

This descriptive cross-sectional study explored practitioner factors influencing pediatric CL uptake in children with significant myopia. Clinics providing pediatric CL services were identified through a national mapping exercise with the Optometry Association of Kenya, covering urban and peri-urban areas.

Of 43 clinics contacted, 38 agreed to participate. Participants included ECPs and children aged 8–18 years with myopia of ≤–0.50 D and astigmatism of ≤–2.75 D. ECPs were eligible if their practice had fitted at least one child with CLs in the preceding two months. Exclusions included patients with ocular prosthesis, ocular comorbidities, therapeutic CL use, high astigmatism (≥–3.00 D), prior unrelated surgery and the absence of a legally authorized representative to provide child consent.

The study defined its primary outcome as pediatric contact lens uptake, measured by whether a child or adolescent with clinically eligible myopia received or was fitted with contact lenses during the same clinical episode. This binary outcome served as a pragmatic indicator of practitioner influence on pediatric contact lens uptake in routine clinical practice. Exposures and predictors included practitioner-reported factors such as child motivation to wear lenses, trial-lens availability, perceived self-esteem benefits among others. Potential confounders comprised practitioner demographics and clinic-level characteristics.

### 2.2 Sample size determination

Sample size was calculated a priori with G*Power v3.1.9.7 (Heinrich Heine Universität Düsseldorf, Germany) using multiple linear regression with 12 predictors of contact lens uptake25. A pilot model explained 13–17 % of variance, so a medium effect size (f^2^ = 0.15) was applied. With α = 0.05 and power (1–β) = 0.80, the minimum required sample was 127 participants (numerator df = 12, denominator df = 114, critical F = 1.838). Allowing 10 % for non-response increased the target to 140.

### 2.3 Instrument of data collection

#### 2.3.1 Questionnaire design and pilot study

A structured questionnaire captured demographics, professional experience, practice setting, age thresholds for CL fitting, and attitudes toward pediatric CL use. Validity was tested in a three-phase, two-week pilot: first, content review and revision; second, expert assessment by three senior optometrists for face validity; and third, pretesting at two eye clinics for construct validity. Feedback improved clarity and usability. The final tool comprised demographic items plus tailored Likert scales for clinicians managing fewer or more than two pediatric CL cases per month.

#### 2.3.2 Clinical Record Abstraction

Data on refractive errors and final optical products dispensed to children and adolescents were abstracted from routine clinical records during consultations by ECPs retrospective analysis. The dependence on secondary clinical data, together with the obligation to protect patient privacy, rendered public participation neither feasible nor appropriate within the study design.

### 2.4 Data collection and analysis

Questionnaire responses (n = 165) were entered into Excel 2016, cleaned for entry errors, and securely stored; 148 questionnaires were completed and analyzed (response rate ∼ 90%). Variables were coded as: gender (1 = male, 2 = female), Likert responses (0 = disagree, 1 = neutral, 2 = agree), clinic type (1–5), and age of CL initiation (1 = <8 years to 6 = not appropriate ≤18 years).

Data analyses were conducted in SPSS v25 and R v4.4.2. Descriptive statistics were generated and odds ratios (ORs) with 95 % confidence intervals (CIs) were estimated. Quantitative variables such as years of practice and number of pediatric contact lens fittings per month were analyzed as continuous measures to preserve information and statistical power. A multivariate logistic regression model was fitted, selecting predictors based on theoretical relevance or bivariate p < 0.10. Further modelling employed ridge regression (λ = 0.007) with 10-fold cross-validation and bootstrap resampling to limit over-fitting and enhance estimate robustness.

## 3.0 Results

### 3.1 Social demographic information of study participants

Most ECPs enrolled in this study (n=148) were male (61.5%) and 38.5% female. Mean CL practice duration was 2.79 years, with 89.9% under five years. Qualifications were as follows: 55.4% bachelor’s, 39.9% college-level, and a smaller proportion with master’s degrees (3.4%). Nearly 95.9% lacked CL organization affiliation. Practice settings were mainly franchised outlets (33.1%), standalone clinics (20.9%), and academic institutions (8.1%).

### 3.2 prescribing patterns for pediatric myopia correction among ECPs in Kenya

A marked preference for early spectacle fitting was observed, with (79.1%) of ECPs recommending spectacles as the initial form of correction for children under eight years (Table 2). In contrast, contact lens fitting was generally deferred, with nearly three-quarters of respondents (72.3%) reporting that they initiate CL prescriptions from age eight onward. The modal age for introducing CLs following spectacles correction was 10–12 years (39.2), whereas the preferred age for recommending CLs as the primary correction modality was 13–14 years (36.5%).

**Table 1.** Socio-demographic characteristics of study participants.

| Practitioner-related variable | Category | n (%) |
| --- | --- | --- |
| Sex | Male | 91 (61.5) |
|  | Female | 57 (38.5) |
| Paediatric CL <sup>a</sup> patients/month | ≤2 children | 111 (75.0) |
|  | >2 children | 37 (25.0) |
| Years <sup>b</sup> in CL practice | 1–2 years | 64 (43.2) |
|  | 3–4 years | 69 (46.6) |
|  | ≥ 5 years | 15 (10.1) |
| Highest qualification | Vocational/technical | 2 (1.4) |
|  | College | 59 (39.9) |
|  | Bachelor's | 82 (55.4) |
|  | Master's | 5 (3.4) |
| CL <sup>a</sup> -related membership | IACLE member | 5 (3.4) |
|  | FIACLE | 4 (0.7) |
|  | None | 139 (95.9) |
| Primary practice | Practice owner | 16 (10.8) |
|  | Franchise | 49 (33.1) |
|  | Standalone practice | 31 (20.9) |
|  | Hospital-based/multidisciplinary | 40 (27.0) |
|  | Educational institution | 12 (8.1) |
| Children's optical correction | Glasses | 57 (38.5) |
|  | Contact lenses | 43 (29.1) |
|  | No correction | 48 (32.4) |

<sup>a</sup> CL = contact lens; n = frequency; % = percentage share.
<sup>b</sup> Calculated mean = 2.79

**Table 2.**
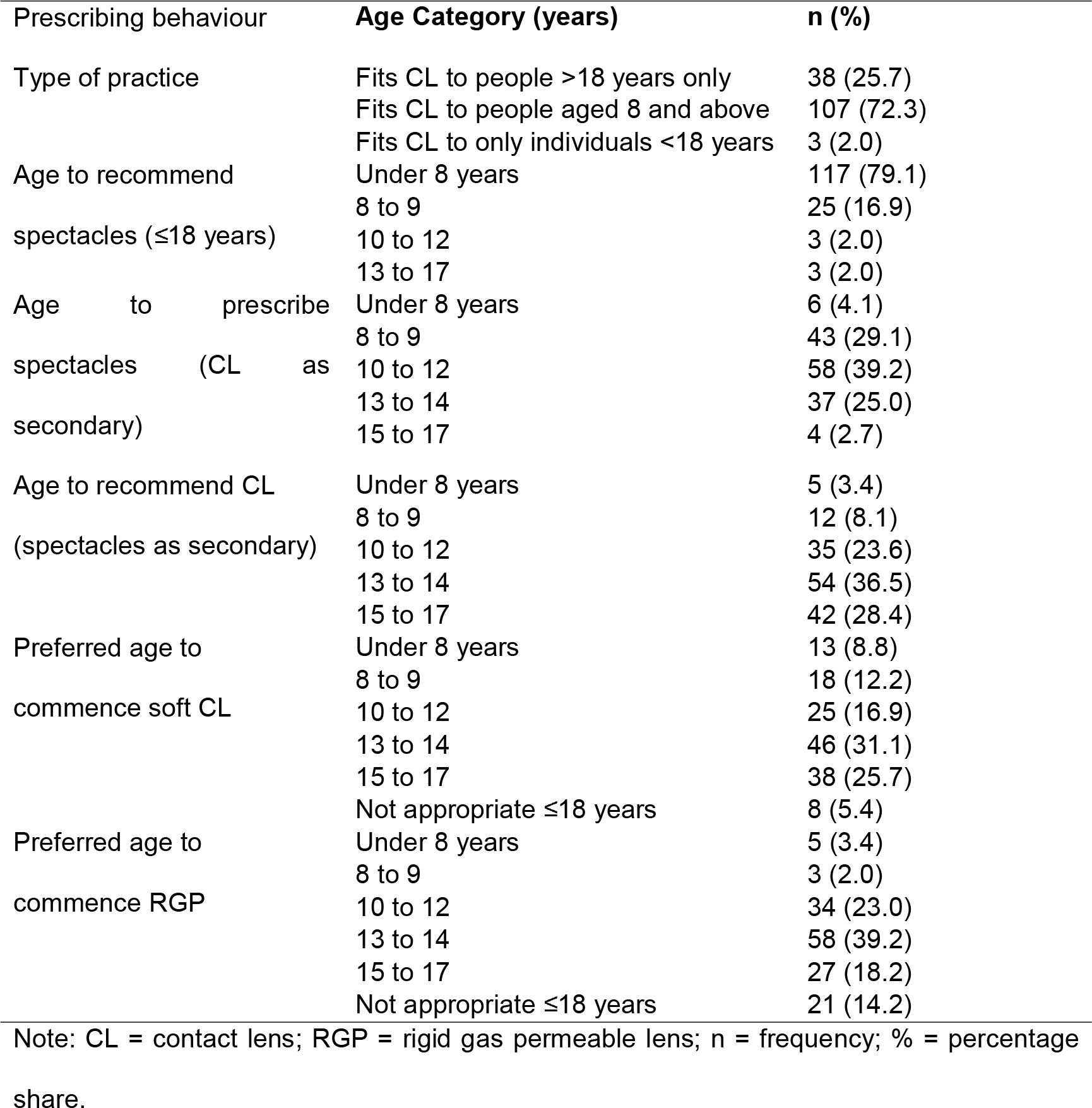
Prescribing patterns for paediatric myopia correction among ECPs.

Regarding specific lens types, 31.1% of ECPs reported initiating soft contact lens fitting at ages 13–14. In contrast, rigid gas-permeable lenses were introduced slightly later, with 39.2% initiating fitting at the same age range and 14.2% considering them unsuitable for patients under 18 years.

### 3.3 Practitioner-perceived enablers of CL uptake in children and adolescents

Table 3 presents practitioner-related factors influencing pediatric contact lens fitting in Kenya. Trial lens use was the leading enabler, endorsed by 62.8% of respondents. Proactive discussion of CL options with families was similarly important (60.1%), although 14.9% remained uncertain. Child motivation (61.5%) and practitioner-perceived self-esteem benefits for children (56.1%) underscored the importance ECPs placed on psychosocial factors in pediatric CL fitting, with few neutral responses (≤10%). Views on patient selection were divided, with 50.0% agreeing that selecting the “right” patients improves outcomes and 49.3% disagreeing. Overall, 52% viewed pediatric CLs as a market opportunity.

**Table 3.** Perceived factors promoting contact lens uptake among children and adolescents with myopia.

| Index <sup>a</sup> | No idea | Disagree | Agree |
| --- | --- | --- | --- |
|  | n (%) | n (%) | n (%) |
| Confidence boost from trial lenses | 11 (7.4) | 44 (29.7) | 93 (62.8) |
| Child's motivation to wear CL □ | 2 (1.4) | 55 (37.2) | 91 (61.5) |
| Perceived CL □ benefits to self-esteem | 10 (6.8) | 55 (37.2) | 83 (56.1) |
| Proactive communication with patients | 22 (14.9) | 37 (25.0) | 89 (60.1) |
| Strategic patient selection | 1 (0.7) | 73 (49.3) | 74 (50.0) |
| Perceived market opportunity | 11 (7.4) | 60 (40.5) | 77 (52.0) |

□ Multiple responses allowed.
□ CL = contact lens; n = frequency; % = percentage share.

### 3.4 Perceived practitioner-related factors hindering CL uptake children and adolescents

Table 4 summarizes ECPs’ perceived barriers to pediatric contact lens uptake in Kenya. Limited public awareness emerged as the most frequently cited challenge, with 62.8% of practitioners agreeing that it delayed consideration of CLs. Parental control was also influential, reported by 51.4% as a hindrance to adoption. Views on knowledge and pediatric fitting skills were divided, with 46.6% agreeing and 43.9% disagreeing that these posed challenges. Low confidence appeared more prominent—acknowledged by 54.1% compared to 33.1% who disagreed. Access to trial lenses presented a practical constraint, with opinions nearly evenly split (44.6% versus 43.2%). Meanwhile, communication challenges were endorsed by 39.9%, while 52.7% disagreed, suggesting that practitioners viewed systemic barriers, rather than interpersonal factors, as the major barriers.

**Table 4.** ECP-perceived barrier factors hindering contact lens uptake among children and adolescents with myopia.

| Index a | No idea | Disagree | Agree |
| --- | --- | --- | --- |
|  | n (%) | n (%) | n (%) |
| Trial lenses are not readily available | 18 (12.2) | 64 (43.2) | 66 (44.6) |
| Parental controls | 7 (4.7) | 65 (43.9) | 76 (51.4) |
| Lack of knowledge in fitting paediatric CL □ | 14 (9.5) | 65 (43.9) | 69 (46.6) |
| Low CL □ awareness among public | 9 (6.1) | 46 (31.1) | 93 (62.8) |
| A communication barrier when it comes to convincing new CL □ users | 11 (7.4) | 78 (52.7) | 59 (39.9) |
| Not confident enough in fitting CL □ | 19 (12.8) | 49 (33.1) | 80 (54.1) |

□ Multiple responses allowed.
□ CL = contact lens; n = frequency; % = percentage share.

### 3.5 Extent of influence of ECPs factors on the uptake of CL in children and teenagers

Multivariate logistic regression (Table 5) showed extent of influence of ECP perceived factors to CL uptake in children and adolescents with significant myopia in Kenya. The highest effects were ECP recognition of self-esteem benefits of CL wear (OR 36.24, CI 18.18–63.57, p = 0.01) and child motivation (OR 25.04, CI 4.36–46.03, p = 0.01). Careful patient selection (OR 23.43, CI 12.54–46.28, p = 0.01), proactive ECP-patient communication (OR 9.62, CI 3.58–16.14, p = 0.02) also promoted uptake. Viewing pediatric CL services as a market opportunity further strengthened access (OR 8.74, CI 4.32–14.70, p = 0.01).

**Table 5.** Extent of influence of ECP-perceived factors on CL uptake of children and adolescents with myopia.

adolescents with myopia
|  | Beta | Z | P | OR <sup>c</sup> | 95% CI <sup>c</sup> |  |
| --- | --- | --- | --- | --- | --- | --- |
|  |  |  |  |  | Lower Bound | Upper Bound |
| **ECP - perceived indices to CL uptake <sup>a</sup>** | **Coef.** | **statistic <sup>b</sup>** | **Value <sup>b</sup>** |  |  |  |
| Confidence Boost from Trial Lenses | 1.06 | 2.94 | 0.02 | 2.89 | 1.05 | 5.47 |
| Child's Motivation to Wear CL | 3.22 | 5.69 | 0.01 | 25.04 | 4.36 | 46.03 |
| Perceived CL Benefits to Self-Esteem | 3.59 | 11.14 | 0.01 | 36.24 | 18.18 | 63.57 |
| Proactive Communication with Patients | 2.26 | 6.42 | 0.02 | 9.62 | 3.58 | 16.14 |
| Strategic Patient Selection | 3.15 | 8.98 | 0.01 | 23.43 | 12.54 | 46.28 |
| Perceived Market Opportunity | 2.17 | 6.83 | 0.01 | 8.74 | 4.32 | 14.7 |
| Low availability of Trial Lenses | -1.67 | -4.08 | 0.01 | 0.19 | 0.07 | 0.41 |
| Parental controls | -1.77 | -5.23 | 0.01 | 0.17 | 0.08 | 0.35 |
| Knowledge and Skills Deficiency | -0.60 | -1.30 | 0.19 | 0.55 | 0.21 | 1.51 |
| Low CL Awareness Among Public | -1.93 | -4.50 | 0.01 | 0.14 | 0.05 | 0.32 |
| Communication Challenges | -1.27 | -4.66 | 0.02 | 0.28 | 0.19 | 0.52 |
| Lack of Practitioner Confidence | -1.60 | -4.59 | 0.01 | 0.20 | 0.11 | 0.45 |

□ Reference category: no CL uptake.
□ Multivariate logistic regression at $\alpha = 0.05$ .
□ OR = odds ratio; CI = confidence interval.

Key barriers included ECP-perceived low public awareness of CL (OR 0.14, CI 0.05– 0.32, p = 0.01), limited trial lenses (OR 0.19, CI 0.07–0.41, p = 0.01), ECP-related communication challenges (OR 0.28, CI 0.19–0.52, p = 0.02), and low practitioner confidence (OR 0.20, CI 0.11–0.45, p = 0.01). parental reluctance was also significant (OR 0.17, CI 0.08–0.35, p = 0.01), while Self-reported knowledge gaps were not (OR 0.55, p = 0.19).

## 4.0 Discussion

### 4.1 Sociodemographic

This study profiles Kenyan CL ECPs, where males made up nearly two-thirds of the respondents in the study. Most were early in their careers, averaging 2.8 years of contact-lens practice, and few had more than five years’ experience. Such limited exposure likely reinforces cautious pediatric fittings. Over half held bachelor’s degrees, about a quarter reported only college-level training, and few possessed a master’s degree. Nearly all ECPS lacked membership in professional contact-lens organizations such as IACLE, limiting access to structured continuing education.

Practice settings were unevenly distributed, with most practitioners in commercial outlets—franchises (33.1%) and standalone clinics (20.9%)—while only 8.1% worked in academic or training facilities. These commercial environments broaden service reach but may often prioritize transactional refractive care over comprehensive pediatric eye management, limiting access [20].

International studies show many practitioners lack confidence in recommending CLs for children [21]. In Kenya, limited pediatric lens access, low awareness, and few specialist ECPs compound these barriers [19].

### 4.2 ECPs prescribing patterns in pediatrics and adolescents with significant myopia

Age-based preferences were clear: nearly three-quarters recommended spectacles first in late childhood, reserving CLs primarily for early to mid-teen years. Global research reveals significant intra-regional differences in pediatric myopia management4. In the UK, 79% of ECPs avoided myopia control [3], highlighting the need to foster evidence-based pediatric myopia management practices [22–24].

### 4.3 Motivators to contact lens fitting in pediatric and adolescents: Practitioner and systemic factors

Several factors shaped pediatric CL prescribing in Kenya. Psychosocial benefits— particularly beliefs that lenses improve children’s self-esteem—were the strongest predictor of prescribing, consistent with the ACHIEVE trial and local reports of enhanced self-image and social acceptance [12]. Perceptions of these benefits varied by practitioner experience and patient context [15, 24, 25], highlighting that social determinants influence myopia-management decisions as much as clinical evidence.

Child motivation strongly increased prescribing, aligning with global findings that motivated young wearers show higher satisfaction and adherence [26, 27]. This both predicts better outcomes and reflects respect for children’s role in care decisions.

Communication emerged as another vital facilitator. Practitioners who initiated discussions about contact lenses were significantly more likely to prescribe, supporting evidence that early dialogue helps normalize lens wear and facilitates family decision-making [15, 25]. Offering trial lenses—though modest—further increased uptake by reducing anxiety and building confidence, underscoring the importance of patient-centered communication.

Clinicians were divided on selective patient screening: about half endorsed targeted selection, while others warned that excessive gatekeeping can delay access to effective myopia control. This highlights a broader tension between safeguarding clinical outcomes and ensuring equitable access to innovative interventions [16].

Finally, clinics where ECPs viewed contact lens services as a market opportunity were more likely to fit children. Similar trends have been observed in China, where commercial framing can expand access [28], although Australian surveys warn that profit motives may influence clinical standards.

### 4.4 Barriers to contact lens fitting in pediatric and adolescents: Practitioner, parental, and systemic factors

Children and adolescents were markedly less likely to receive contact lenses when multiple barriers coexisted. Low public awareness was a major limiting factor, consistent with findings from Ghana—where over one-quarter avoided CL use due to poor information [22] —and Kenya, where 96.7% reported no awareness of CLs [29]. Without targeted education, potential users rarely request lenses even when clinically indicated.

A clear gap emerged between self-reported skills and actual prescribing. Although technical competence was not a significant predictor, low confidence in pediatric CL fitting strongly associated with reduced prescribing. Many practitioners considered themselves capable but hesitated in practice, likely due to limited clinical exposure (mean 2.79 years) and strategic communication challenges with parents and children. Prior studies[30, 31] similarly report that early-career optometrists feel underprepared for complex pediatric fittings.

Poor practitioner–patient communication also independently predicted reduced prescribing, aligning with prior work showing that weaker dialogue lowers CL adoption[32, 33]. Financial concerns may exacerbate this gap: for instance, a Chinese study found that 13.8% of ECPs avoided discussing costly treatments to patients for fear of being perceived as commercially biased [34], suggesting similar hesitancy in resource-limited settings like Kenya.

Systemic barriers identified were also significant. Scarcity of trial lenses had a strong association with limited uptake of services in the study. ECPs who identified it as a barrier were often unable to demonstrate fit or comfort to families—a step critical in building parental trust.

Finally perceived parental controls to CL use was widely reported, with analysis showing it significantly associated with reduced prescribing of pediatric myopia CLs by Kenyan ECPs. Previous Kenyan and Chinese studies have shown that parents often delay CL adoption due to age-related concerns, and personal experiences [19, 34].

All together these current results indicate parental control, awareness, and system-level enablers carry greater weight in actual prescribing behaviors in pediatric contact lens fitting in Kenya. Uptake is supported by trial lens availability, proactive communication, and psychosocial benefits such as improved self-esteem. Key barriers include low public awareness, parental misconceptions, limited practitioner confidence, and poor communication. Addressing these challenges requires practitioner training, public education, early-career support, and collaborations such as IACLE. Expanding access to trial lenses through subsidies and developing evidence-based protocols can normalize pediatric CL use and improve visual outcomes.

The findings are generalizable to urban and peri-urban optometric settings in Kenya and similar low-resource contexts with emerging pediatric contact lens services. Inclusion of multiple clinics enhances representativeness, though generalizability to rural or different regulatory environments is limited. Despite cross-sectional design constraints, results offer transferable insights into practitioner and system-level determinants of pediatric myopia care.

#### Literature Search

A systematic search was conducted in PubMed, Embase, Web of Science, and Scopus from February 2024 to 2025, for studies published between 2000 and 2025. Search terms combined subject headings and keywords (e.g., *pediatric myopia, contact lens fitting, eye care practitioners, barriers, motivators, refractive correction*) using Boolean operators.

## Acknowledgements

We thank the participating eye care practitioners for their support in data collection.

## Ethics Approval

Approved by Masinde Muliro University of Science and Technology (MMUST/IERC/029/2021) and the National Commission for Science, Technology, and Innovation (NACOSTI/P/21/14780).

## Declaration

This manuscript is original, not under review elsewhere, and all authors have approved the final version.

## Consent

Written informed consent and/or assent was obtained from all participants.

## Use of Generative AI

ChatGPT-4™ was used for grammar and clarity enhancement. All content was reviewed and verified by the authors.

## Data Availability

All anonymized data and analytical workflows are publicly available via Figshare:

Kololi G, Sum TJ. Barriers and motivators to pediatric contact lens fitting. 2025;1(1):1. https://doi.org/10.6084/m9.figshare.30479246.v2

